# HRD-One: CLINICAL VALIDATION AND PERFORMANCE ASSESSMENT. Comparison between Myriad’s myChoice®, SOPHiA GENETICS® SOPHiA Homologous Recombination Solution and AmoyDx® HRD Focus Panel

**DOI:** 10.1101/2023.03.28.21264560

**Authors:** Rodrigo Guarischi-Sousa, José Eduardo Kroll, Adriano Bonaldi, Paulo Marques Pierry, Luiz Gustavo de Almeida, Camila Alves Souza, Juliana Santos Silva, Darine Villela, Fabiana Marcelino Meliso, Maria Fernanda Grillo Milanezi, Cristovam Scapulatempo Neto, Guilherme Lopes Yamamoto

## Abstract

Homologous Recombination Repair (HRR) testing has become increasingly important in clinical genomic labs due to the use of poly-ADP-ribose polymerase (PARP) inhibitor therapy for epithelial ovarian, fallopian tube, or peritoneum cancer. While sequencing and copy number variation analysis can identify patients with a pathogenic mutation in BRCA1 or BRCA2 who can benefit from PARPi therapy, there are also patients who may benefit but do not have these mutations. To address this, our lab has developed a test called HRD-One, in partnership with SOPHiA GENETICS, that can detect sequence variants in genes involved in HRR, as well as genomic scars that indicate Homologous Recombination Deficiency (HRD), which may be present even when a pathogenic variant is not detected. We tested 59 high-grade serous epithelial ovarian cancer samples using HRD-One and found that it had an overall categorical concordance of 94.74% with Myriad’s myChoice® score, which is a commercial HRD test. 12 out of 13 samples that carried a pathogenic or likely pathogenic variant in BRCA1/2 also had a positive HRD-One score, and 9 samples in which a pathogenic variant in BRCA1/2 was not identified had a positive score in both HRD-One and myChoice®. Of the samples that passed quality control, we observed an average of 1.62 points variation between replicates on a scale from -25.0 to +25.0. We also found that low-confidence results were associated with a low DNA input and the age of FFPE blocks, while the estimated tumor percentage in the block, NGS library yield, and score of genomic instability did not have a significant association. We determined that blocks older than 3 years or with a DNA input of less than 25ng are not reliable for producing high-quality results. Finally, we validated the HRD-One test with SOPHiA Homologous Recombination Solution (Library Prep kit II) and correlated it to myChoice®, and found that the AmoyDx® HRD Focus Panel had the same sensitivity but a higher number of false positive samples and therefore lower specificity. Overall, we have shown that HRD-One can provide a reproducible and concordant score for inferring HRD, and an HRD-One score of 2.0 or greater predicts HRD and correlates to Myriad’s myChoice® score of 42 in high-grade serous epithelial ovarian cancers samples that meet our minimum quality criteria.

## INTRODUCTION

Homologous Recombination Repair (HRR) represents an important repair pathway of potentially lethal DNA double strand breaks; its deficiency sets off a phenotype of tumor cells that leads to the accumulation of genetic damage. However, when a second deficiency in DNA replication/repairing caused by the lack or inhibition of poly-ADP-ribose polymerase (PARP) is present, instead of benefiting cancer cells, a mechanism of synthetic lethality can be achieved in which only cells that lack HRR die when PARP is also inhibited. With the advent of PARP inhibitor therapy for epithelial ovarian, fallopian tube or peritoneum cancer, testing of HRR aberrations as a biomarker of response to therapy was brought to the routine workflow of genomic laboratories. Several mechanisms can cause Homologous Recombination Deficiency (HRD), such as mutations of genes involved in the HRR pathway, specially *BRCA1* and *BRCA2*, methylation of gene promoters, and other unknown mechanisms. The most utilized method for determining the presence of HRD is to sequence known HRR genes and look for deleterious mutations. Another method is to look for the effect (or phenotype) of loss of HRR function by quantifying the degree of genomic aberrations or scars in the genome; this is also called “genomic instability”. Whole Genome Sequencing (WGS) coupled with specialized algorithms trained to recognize patterns characteristic of genomic instability can be used to quantify HRD aberrations. Here we present the clinical validation of HRD-One, an HRD test based on low-pass WGS (lpWGS) data combined with targeted sequencing of HRR genes on tumor samples, and its correlation with a commercial test used for HRD assessment.

## MATERIALS AND METHODS

### Tumor Samples and DNA extraction

Fifty-nine high-grade serous epithelial ovarian cancers samples were retrieved from pathology archives of a private diagnostic laboratory in Brazil. The percentage of neoplastic cells in the total amount of tissue section and in the marked area were described for all samples. The 59 samples were also sent to a reference laboratory for homologous recombination deficiency assessment, for validation purposes. Other 21 FFPE carcinoma samples from distinct primary sites, including ovary, prostate, breast, pancreas, were added to the validation as controls for the evaluation of reproducibility and repeatability, but no known HRD status is available for these cases. This initial validation was performed with SOPHiA Homologous Recombination Solution DNA Library Prep kit II (LPII).Thirty-two of the initial fifty-nine samples, plus another twelve different high-grade serous epithelial ovarian cancers samples that underwent myChoice® testing were used to validate concordance of AmoyDx® HRD Focus Panel with both myChoice® and HRD-One SOPHiA DNA Library Prep kit II. Some samples differ between comparisons due to insufficient sample/DNA quantity to perform all assays with the same exact samples. DNA extraction was performed according to standard laboratory procedures, and concentration determined based on a standard curve generated by serial dilutions of the Control DNA CEPH 1347-02 (Thermo Fisher Scientific), with concentration values ranging from 50 ng/μL to 0,04 ng/μL. gDNA integrity was accessed using the Genomic DNA ScreenTape in a TapeStation equipment (Agilent Technologies).

### Library preparation, sequencing method and data analysis

For the initial validation with 59 samples, DNA libraries were built according to SOPHiA Homologous Recombination Solution protocol (version PM_T1_T2_T3_5.1.96_r1en, November 2019), whereas for the other forty-four samples we tested the AmoyDx® HRD Focus Panel protocol, using the version B1.2 (March 2021). In HRD-One (SOPHiA LPII) validation, the complete analysis of genomic instability involves both genome sequencing at a low depth (approximately 1x to 3x coverage), on Illumina sequencing platforms, and evaluation of the presence of pathogenic variants in the genes present in the Homologous Recombination Repair (HRR) genes panel, which includes a total of 16 genes, but for this study we evaluated only *BRCA*. Genomic integrity index (GII) score was calculated based on a deep learning algorithm and was performed on SOPHiA DDM™ platform. For the AmoyDx validation, the patient HRD status is determined by identifying mutations in *BRCA* and determining a genomic scar score using the platform of data analysis from the manufacture (ANDAS System, China).

## RESULTS

### HRD-One VALIDATION (SOPHiA LPII vs myChoice®)

#### Description of validation samples

Table 1 contains a brief description of samples used on this validation as well as the main clinical characteristics of each one, such as: cancer site, estimated tumor content (ETC) and DNA input, which is an important quality parameter directly impacting the feasibility of preparing DNA libraries used for NGS sequencing.

**Table 1.** Description of samples used to validate HRD-One test (SOPHiA LPII).

| Sample | Run | Cancer site | Estimated tumor content (%) | DNA input (ng) | Sample | Run | Cancer site | Estimated tumor content (%) | DNA input (ng) |
| --- | --- | --- | --- | --- | --- | --- | --- | --- | --- |
| 1 | 1 | Ovary | 80 | 150 | 31 | 3 | Ovary | 85 | 100 |
| 2 | 2 | Ovary | 50 | 150 | 32 | 3 | Peritoneum | 90 | 50 |
| 3 | 2 | Ovary | 60 | 150 | 33 | 3 | Ovary | 90 | 50 |
| 4 | 2 | Ovary | 70 | 150 | 10 | 3 | Ovary | 90 | 100 |
| 5 | 2 | Ovary | 70 | 150 | 34 | 3 | Ovary | 95 | 100 |
| 6 | 2 | Peritoneum | 75 | 150 | 35 | 3 | Fallopian tube | 98 | 25 |
| 7 | 2 | Peritoneum | 80 | 150 | 36 | 4 | Ovary | 20 | 25 |
| 1 | 2 | Ovary | 80 | 150 | 37 | 4 | Peritoneum | 20 | 50 |
| 8 | 2 | Ovary | 90 | 150 | 38 | 4 | Ovary | 20 | 100 |
| 9 | 2 | Ovary | 90 | 150 | 39 | 4 | Ovary | 20 | 100 |
| 10 | 2 | Ovary | 90 | 150 | 40 | 4 | Ovary | 30 | 100 |
| 10 | 2 | Ovary | 90 | 150 | 41 | 4 | Ovary | 40 | 25 |
| 11 | 2 | Ovary | 98 | 150 | 42 | 4 | Ovary | 40 | 100 |
| 12 | 3 | Ovary | 30 | 100 | 43 | 4 | Peritoneum | 50 | 50 |
| 13 | 3 | Ovary | 40 | 100 | 44 | 4 | Ovary | 50 | 100 |
| 14 | 3 | Ovary | 40 | 100 | 45 | 4 | Ovary | 60 | 3.9 |
| 15 | 3 | Ovary | 50 | 50 | 46 | 4 | Peritoneum | 60 | 100 |
| 16 | 3 | Peritoneum | 50 | 100 | 47 | 4 | Ovary/Peritoneum | 60 | 100 |
| 17 | 3 | Ovary | 60 | 25 | 48 | 4 | Ovary | 60 | 100 |
| 18 | 3 | Ovary | 60 | 100 | 49 | 4 | Ovary | 60 | 100 |
| 19 | 3 | Ovary | 60 | 100 | 50 | 4 | Fallopian tube | 70 | 50 |
| 20 | 3 | Ovary | 60 | 100 | 51 | 4 | Ovary | 70 | 50 |
| 21 | 3 | Ovary | 70 | 100 | 21 | 4 | Ovary | 70 | 100 |
| 22 | 3 | Peritoneum | 70 | 100 | 52 | 4 | Ovary | 80 | 19.2 |
| 23 | 3 | Fallopian tube | 70 | 100 | 53 | 4 | Ovary | 80 | 50 |
| 24 | 3 | Ovary | 80 | 100 | 54 | 4 | Ovary | 80 | 100 |
| 25 | 3 | Ovary | 80 | 100 | 55 | 4 | Ovary | 80 | 100 |
| 26 | 3 | Fallopian tube | 80 | 100 | 56 | 4 | Ovary | 80 | 100 |
| 27 | 3 | Ovary | 80 | 100 | 57 | 4 | Ovary | 90 | 100 |
| 28 | 3 | Ovary | 80 | 100 | 58 | 4 | Ovary | 90 | 100 |
| 29 | 3 | Ovary | 80 | 100 | 59 | 4 | Ovary | 90 | 100 |
| 30 | 3 | Ovary | 80 | 100 |  |  |  |  |  |

#### Discrimination of positive/negative samples

In order to maximize the discrimination power of positive/negative samples, we performed an evaluation of different thresholds to find which value maximizes specificity between positive and negative groups. Based on correlation with Myriad’s myChoice® test, this theoretical threshold was computed to be 2.0, where samples with higher values shall be classified as positive and samples with lower values as negative. Noteworthy, a narrow uncertainty margin between 0.2 and 3.2 was observed, in which discrimination between classes is less clear (Figure 1).

**Figure 1.**
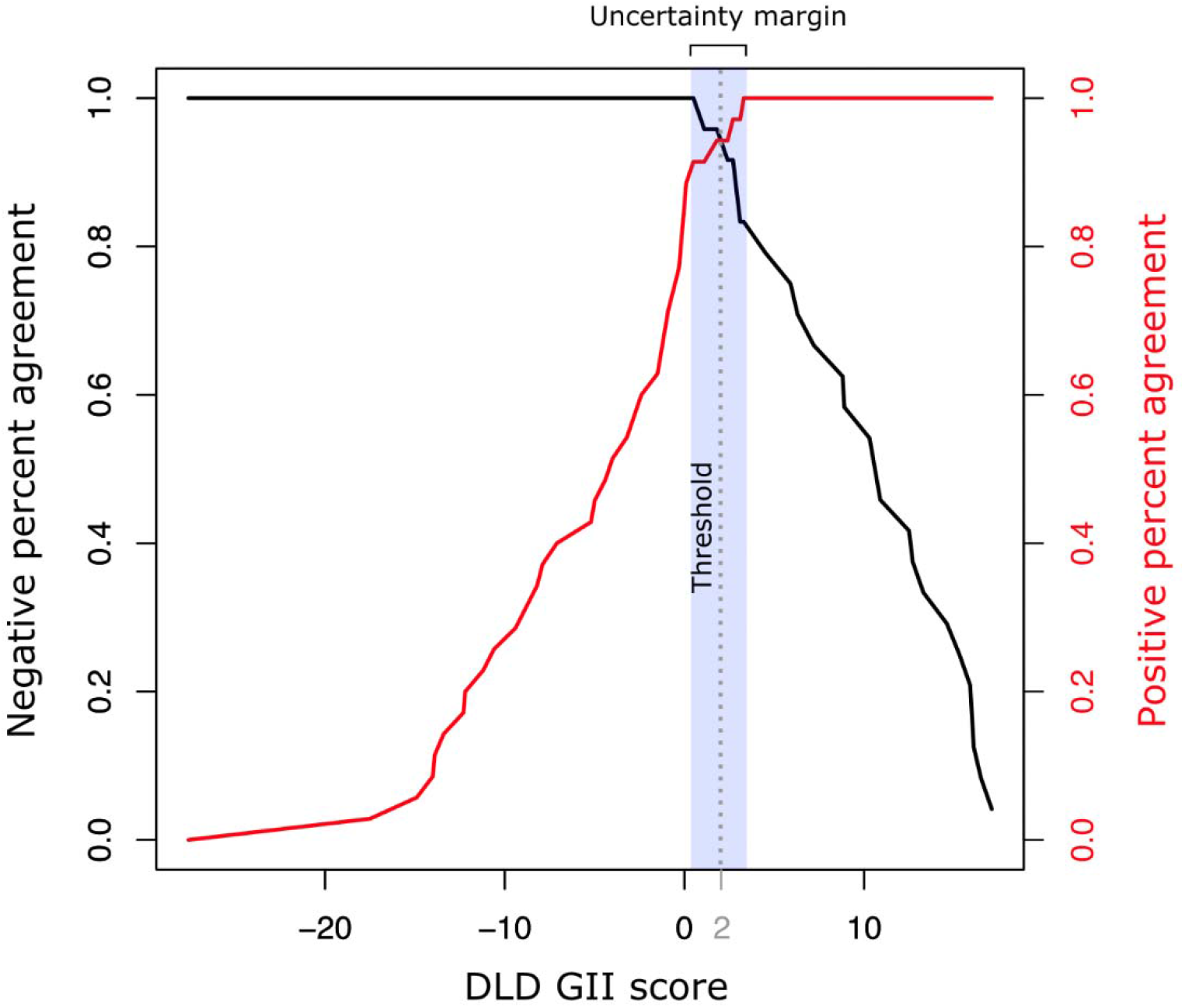
Threshold analysis based on the specificity equilibrium between the positive and negative groups.

#### Correlation between methodologies for genomic instability/integrity detection

An overall strong correlation between known genomic instability scores (myChoice**®** GIS) and DLD GII scores (SOPHiA Homologous Recombination Solution GII) was observed (R=0.87, p-value=2.2×10^−16^; Figure 2A). Noteworthy, samples which deviate most from the central trend line are samples flagged as low-confidence during quality-control steps (Figure 2A). For these samples, low tumor content, low DNA input, high level of DNA degradation or weak genomic integrity signal, which impairs signal-to-noise ratio used in the algorithm, are the likely causes of bigger deviations. Considering a threshold of 2.0 to discriminate between positive and negative samples, a concordance of 94.74% with Myriad’s myChoice® test was observed on this cohort (p-value=1.72×10^−12^, Fisher’s exact test; Figure 2C). The analytical sensitivity of the test was 95.7%, the analytical specificity was 94.1%, the positive predictive value (PPV) was 91.7% and the negative predictive value (NPV) was 97% (also when compared with the available HRD commercial test).

**Figure 2.**
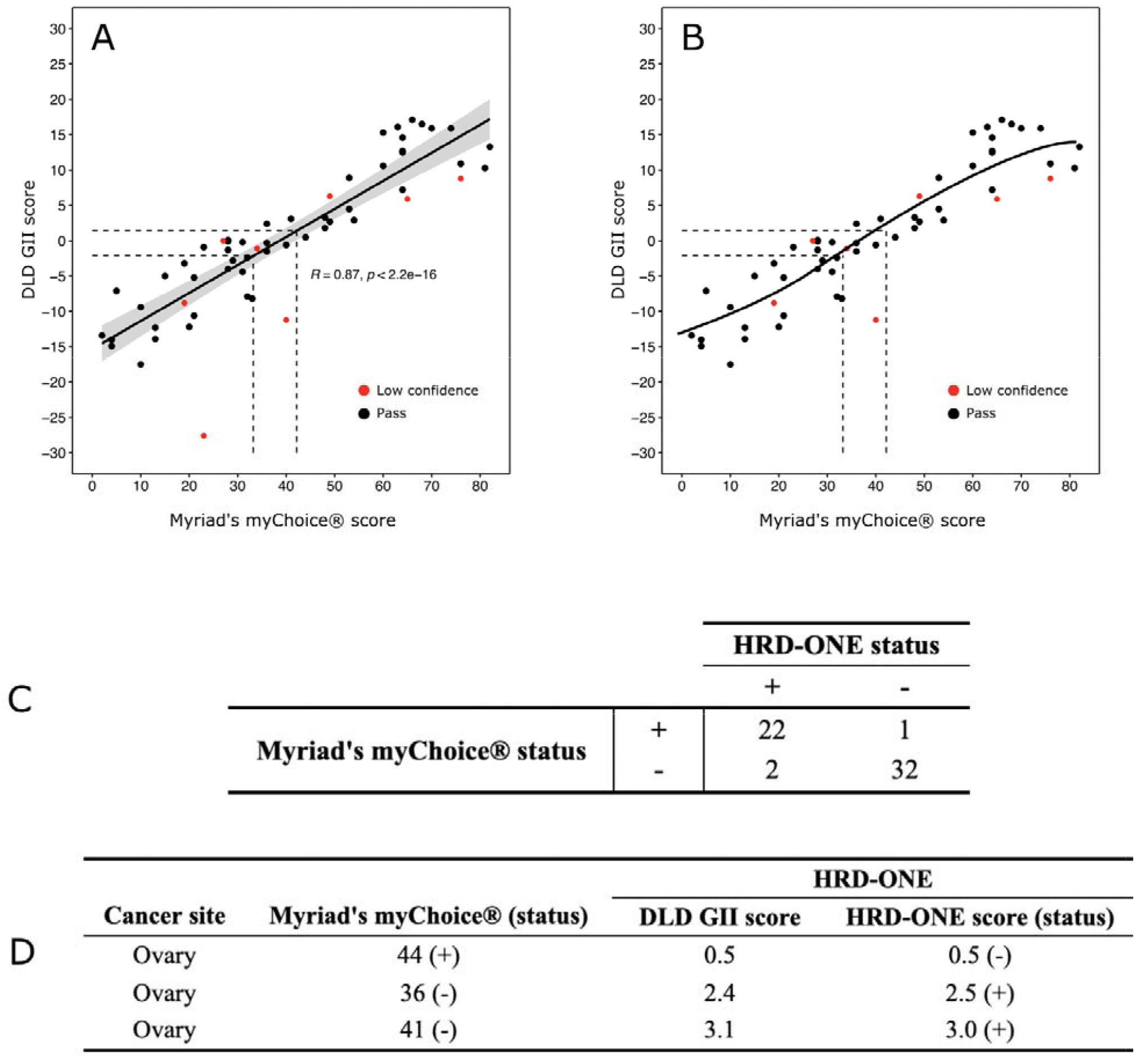
A) A strong Pearson correlation was observed between known GI score and Deep Learning-derived (DLD) GII scores (all samples and replicates); B) Polynomial regression (order 3) of unique samples excluding the outlier; C) Contingency table of positive/negative sample showing concordance between known GII status and HRD-One status; D) Detailed information on the 3 samples with categorical divergencies between known status and HRD-One status.

Three ovarian tumor samples showed categorical discordances between the commercial test and HRD-One (Figure 2D). Two samples with negative status and one with positive status, based on Myriad’s myChoice® test, turned out to be the opposite status by HRD-One, with borderline scores. Considering that reported genomic instability scores from Myriad’s myChoice® were 36, 41 and 44 for these 3 samples, close to the stablished threshold for the current standard test (42), and the uncertainty margin predicted for HRD-One (between 0.2 and 3.2), extra caution is expected when counselling patients with such borderline results.

Both linear and polynomial regressions presented very good correlations (Figure 2A and 2B). Due to the facts that: a) scores below 30 (−3.0) are clearly negative and above 50 (+5.5) are clearly positive; b) the order 3 polynomial regression is almost linear between 30 and 50 and has a slightly better fit than the linear regression; c) the DLD GI score seems to cap at both extremities; we have opted to establish a correlation table utilizing the polynomial regression (Table 2).

**Table 2.**
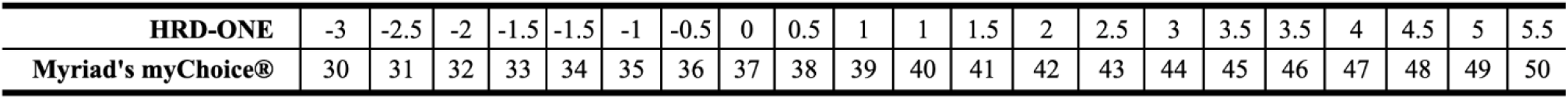
Correlation between HRD-One score and commercial test score.

**Table 3.** The two samples with inconclusive results from benchmark commercial test were also marked as low confidence within our validation set. The likely cause is the hindered capacity of extracting enough DNA input for library preparation.

| Sample | Cancer site | Estimated tumor content | DNA input (ng) | Sample quality | Library yield (ng/μL) | Myriad's myChoice® status | HRD-ONE |  |
| --- | --- | --- | --- | --- | --- | --- | --- | --- |
|  |  |  |  |  |  |  | QA status | DLD GII score |
| 1 | Ovary | 60 | 3.9 | bad | 66.8 | inconclusive | low confidence | -2 |
| 2 | Ovary | 80 | 19.2 | bad | 154.8 | inconclusive | low confidence | -25.4 |

#### Reproducibility and robustness of GII scores

In order to assess the reproducibility of DLD GII score, 6 samples were sequenced as replicas. A very high correlation was observed either within or between sequencing batches for all samples tested, reassuring the reproducibility and robustness of GII scores across multiple experiments (R=1.0; p-value < 4.5×10-6; Figure 3). The mean variability observed between replicas was 1.62

**Figure 3.**
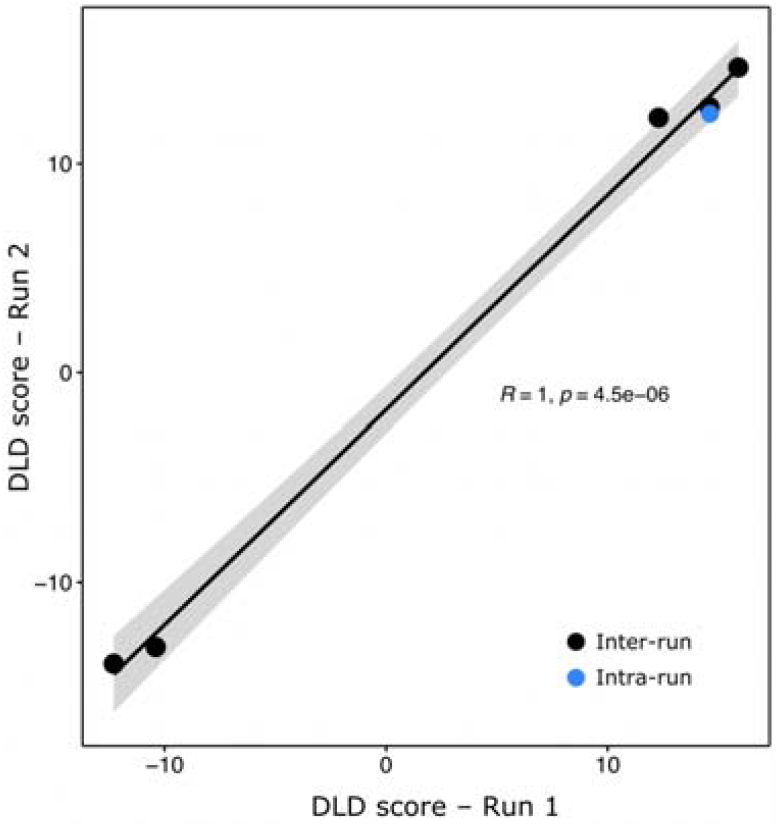
Pearson correlation between DLD GII scores for inter and intra-run replicas.

±0.88. Due to this level of variability and the sake of facilitating interpretation of clinical results, we opted for using scores rounded to half a point to represent the final HRD-One GII score (Table 2).

#### Correlation between HRR and HRD-One score

Among the 59 unique samples 13 presented a pathogenic or likely pathogenic variant in either *BRCA1* or *BRCA2* (22%); and only 1 of the 13 HRR positive samples had a negative HRD score (7,7%), which was also concordant between HRD-One (score -5.0) and Myriad’s myChoice® (score 15). These data are in accordance with similar experiments in the literature and confirms first that some of the HRR positive samples might present a negative HRD GII score and second that the HRD-one test concordance with HRR pathogenic mutations was the exact same as the concordance between Myriad’s myChoice® score and HRR pathogenic mutations. HRD-One scores in the 12 samples that are HRR positive ranged from 2.0 to 17.1 (average 9.9), Myriad’s myChoice® scores ranged from 48 to 82 (average 63).

#### Pre analytical variables associated with low confidence experiments

Especially when dealing with challenging materials such as FFPE samples, pre-analytical steps are important to ensure a reliable analytical result. We sought to investigate a set of parameters and how they associate with low-confidence warnings indicated by the algorithm.

Within our dataset, we observed significant differences between pass and low-confidence cases for DNA input (p-value=4.6×10^−3^; Figure 4A) and age of FFPE blocks (p-value=3.9×10^−2^; Figure 4B). All other pre-analytical variables tested of estimated tumor content (p-value=0.2242; Figure 4C) and library yield (p-value=0.1403; Figure 4D) did not show significant differences between the two classes. Importantly, we could not observe an association between DLD GII scores and low-confidence flags (p-value=0.1353; Figure 4E), reenforcing that no clear bias towards positive or negative cases is expected regarding quality matters.

**Figure 4.**
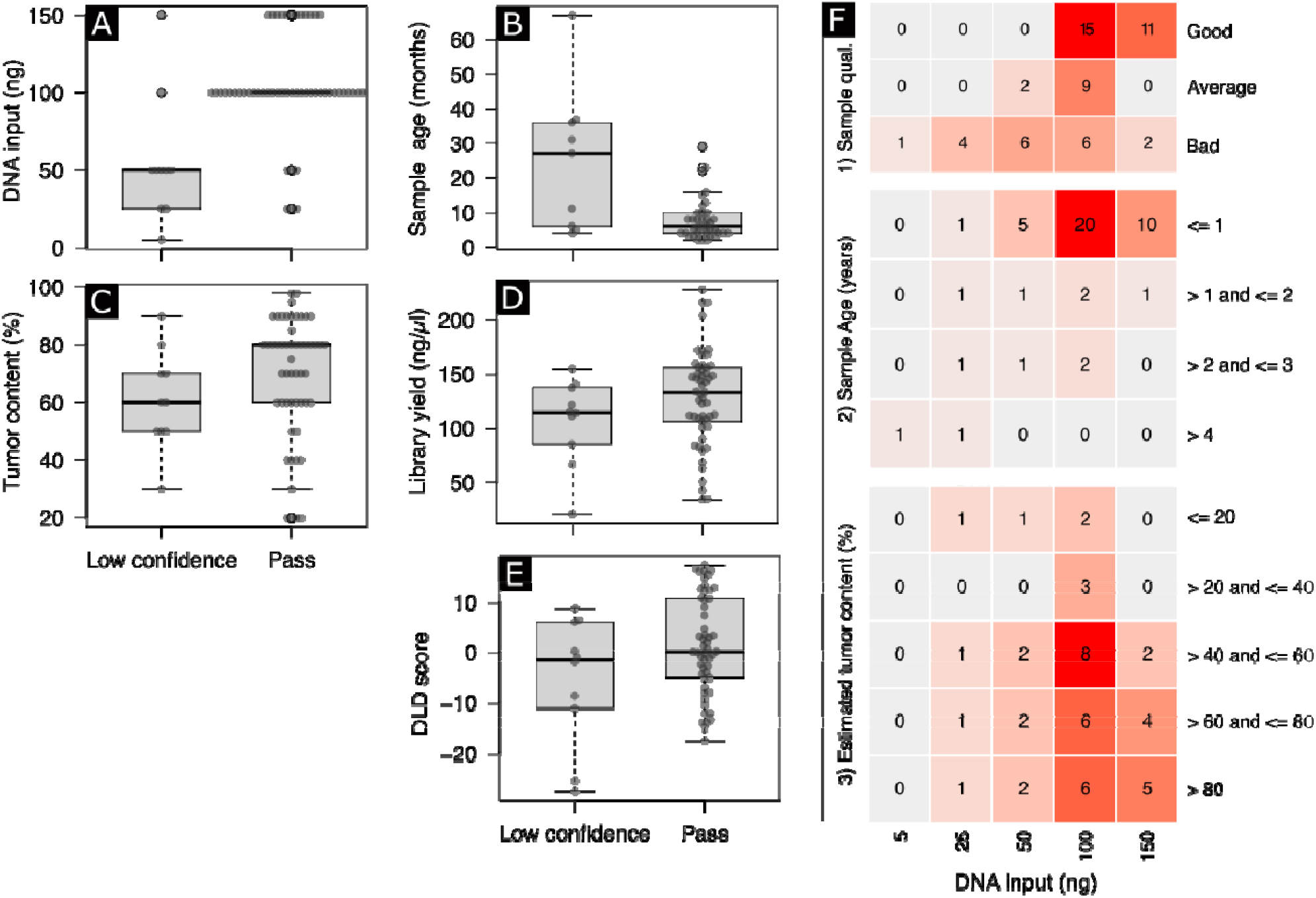
Association between low confidence experiments and major pre-sequencing parameters. Significant association was observed for A) DNA input and B) age of FFPE blocks, while no significant association was observed for C) estimated tumor content, D) library yield and E) DLD GI scores. F) Association between DNA input, the isolated most significant pre-sequencing parameter, with sample quality, age of FFPE blocks and estimated tumor content.

Samples with lower ETC, specially less than 30%, are expected to fail on a higher rate due to a lower signal for the algorithm to calculate genomic integrity (Figure 4C).

Finally, we sought on evaluating the association between DNA input, the isolated most significant pre-sequencing parameter, with sample quality, age of FFPE blocks and ETC. Samples with good gDNA profile tend to yield more while samples with average or poor show a less clear pattern. Similarly, FFPE blocks with less than 1 year tend to yield good amounts of input DNA, blocks between 1 and 3 years show a less clear trend, while blocks with more than 4 years old usually have a hindered extraction. In our validation, we were able to extract more than 100 ng of functional DNA from most of FFPE blocks and set a DNA input of 100 ng per reaction, whenever possible to proceed with library preparation for NGS (Figure 4F). Older samples and samples on the lower end of input DNA (5 to 50 ng) have shown to produce low confidence results, we have therefore adopted 50 ng as the minimum recommended input to proceed with HRD testing. It is possible that low DNA yield is a consequence of age and/or DNA degradation and therefore and indirect measure of these factors.

#### Samples with inconclusive results

In our validation set, we only had two samples with inconclusive results on Myriad’s myChoice® test, these two samples were also flagged as low confidence by the DLD algorithm. Noteworthy, these two samples had the lowest input DNA among all samples in our validation set (Table 2).

## AmoyDx® HRD Focus Panel VALIDATION (AmoyDx® HRD Focus Panel vs SOPHiA LPII and myChoice®)

### Description of validation samples

Table 4 contains a brief description of the extra samples used on this validation as well as the main clinical characteristics of each one, such as: cancer site, estimated tumor content (ETC) and DNA input, which is an important quality parameter directly impacting the feasibility of preparing DNA libraries used for NGS sequencing. A total of 44 samples, 32 from initial validation (samples: 3, 4, 5, 7, 8, 9, 10, 11, 16, 20, 21, 22, 23, 24, 29, 30, 31, 32, 33, 34, 35, 38, 44, 45, 46, 49, 53, 54, 55, 56, 58 and 59) plus 12 novel samples (60-71) were included.

**Table 4.**
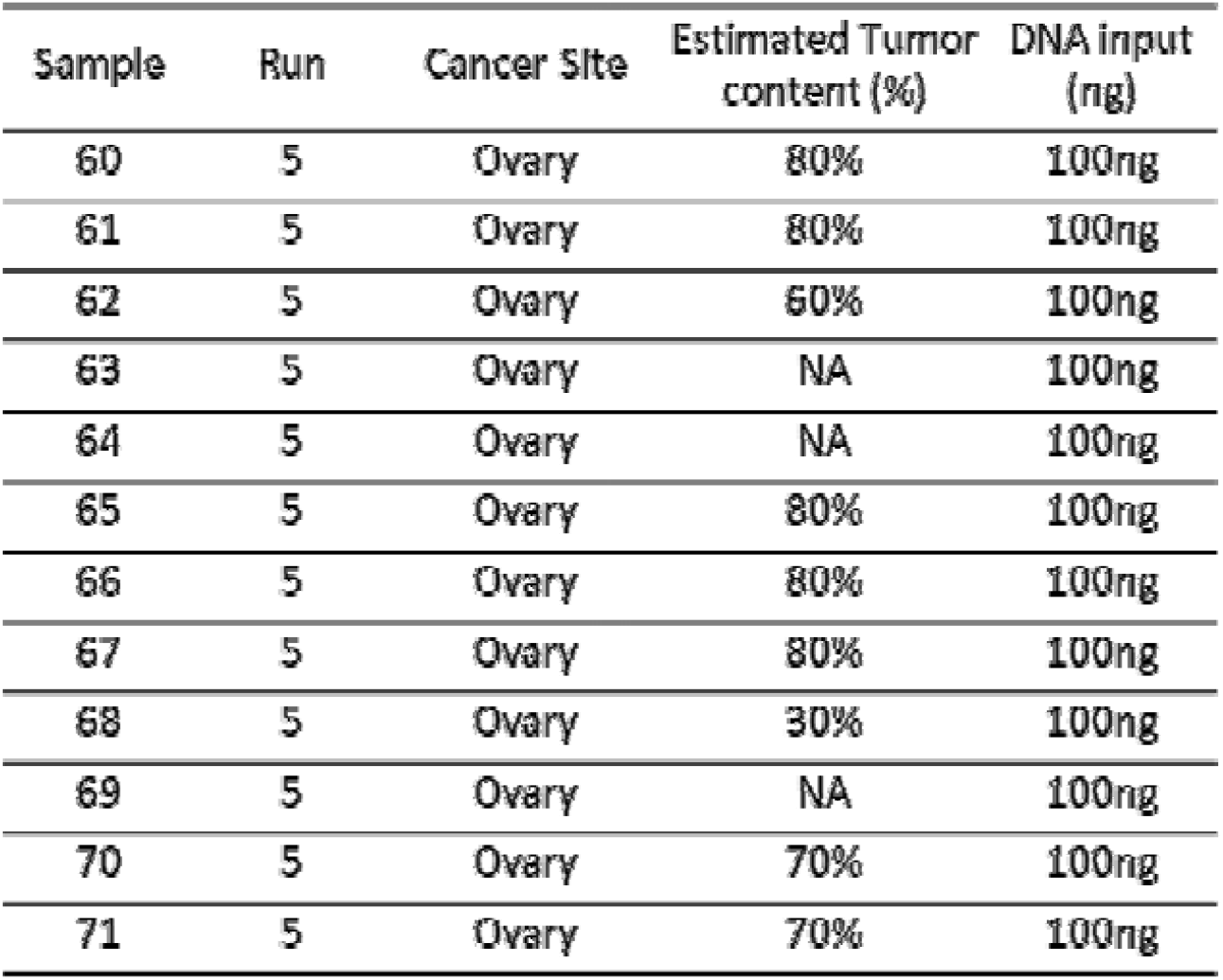
Description of extra samples used to validate AmoyDx® HRD Focus Panel.

Of important note 7 out of 44 samples failed quality control in AmoyDx® HRD Focus Panel (samples: 7, 22, 29, 35, 54, 69 and 71). This proportion is only a bit higher than 7/59 low confidence samples in SOPHiA LPII, but the failed category would result in a necessity for repetition or, in case of lack of enough input material, failed test.

### Correlation between methodologies for genomic instability/integrity detection

Even though there was a strong correlation between known genomic instability scores (myChoice**®** GIS) and GSS (AmoyDx® Genomic Scars Score), the correlation observed for AmoyDx**®** test was lower than the observerd for SOPHiA LPII (Figure 5 A-J). Considering the proposed threshold of 50 to discriminate between positive and negative samples, a concordance of 86.36% with Myriad’s myChoice® test was observed on this cohort (p-value= 1.47 ×10^−7^, Fisher’s exact test; Figure 6A). The analytical sensitivity of the test was 100%, the analytical specificity was 73.9%, the positive predictive value (PPV) was 77.8% and the negative predictive value (NPV) was 100% (also when compared with Myriad’s myChoice®).

**Figure 5.**
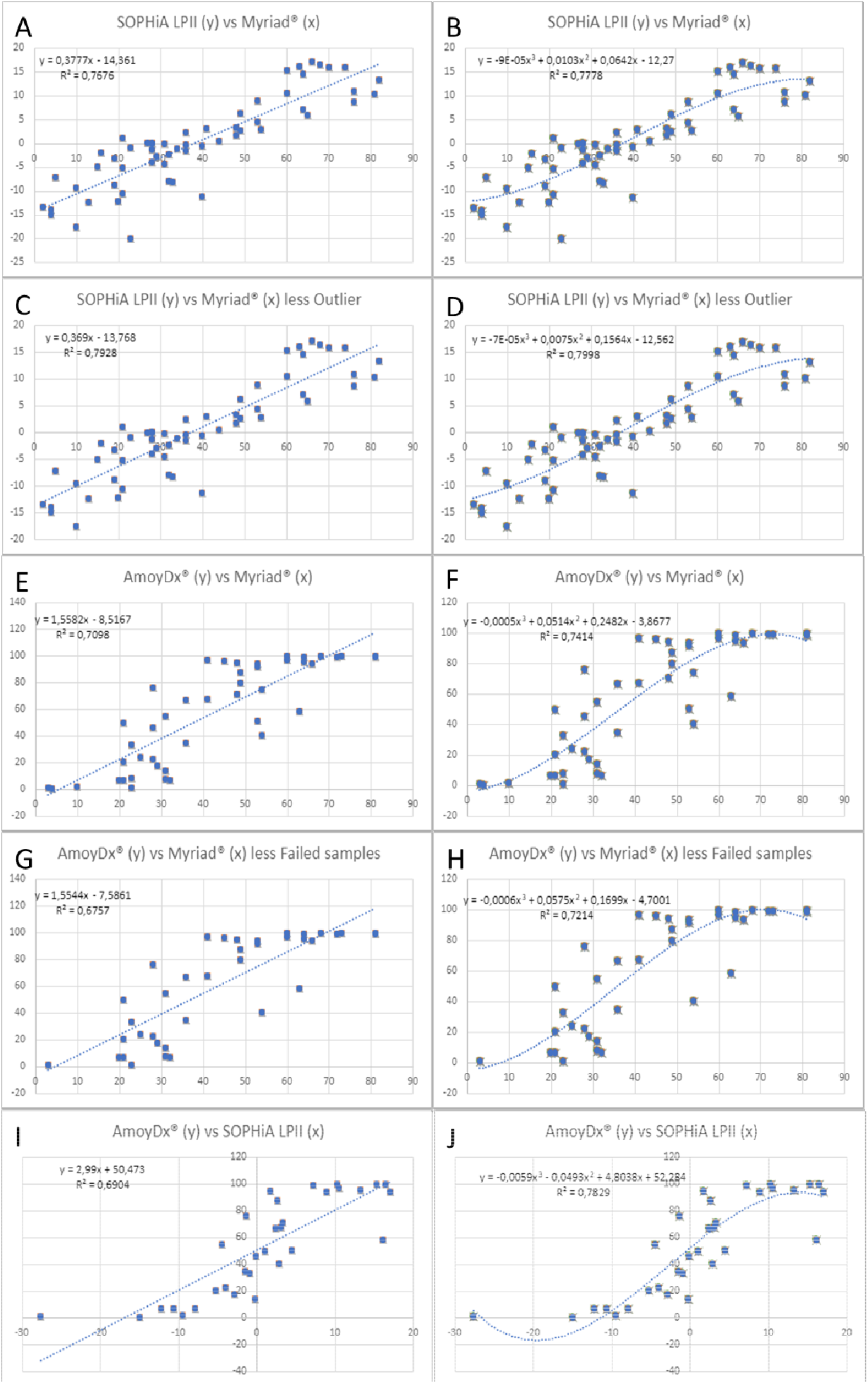
A and B) Linear and polynomial regression (order 3) of unique samples between Myriad’s myChoice® and SOPHiA GENETICS® SOPHiA Homologous Recombination Solution, note polynomial regression R^2^ of 0,78; C and D) Linear and polynomial regression (order 3) of unique samples excluding the outlier between Myriad’s myChoice® and SOPHiA GENETICS® SOPHiA Homologous Recombination Solution, note polynomial regression R^2^ of 0,80; E and F) Linear and polynomial regression (order 3) of unique samples between Myriad’s myChoice® and AmoyDx® HRD Focus Panel, note that polynomial regression R^2^ of 0,74 is lower than the previous two; G and H) Linear and polynomial regression (order 3) of unique samples excluding failed quality samples between Myriad’s myChoice® and AmoyDx® HRD Focus Panel, note that even removing failed samples the polynomial regression R^2^ of 0,72 is lower than all others; I and J) Linear and polynomial regression (order 3) of unique samples between and AmoyDx® HRD Focus Panel and SOPHiA GENETICS® SOPHiA Homologous Recombination Solution, note that, even though AmoyDx® solution has lower correlation with Myriad’s myChoice®, it has high correlation (polynomial regression R^2^ of 0,78) with SOPHiA GENETICS® solution, possibly due to similarities in the calculation methods.

**Figure 6.**
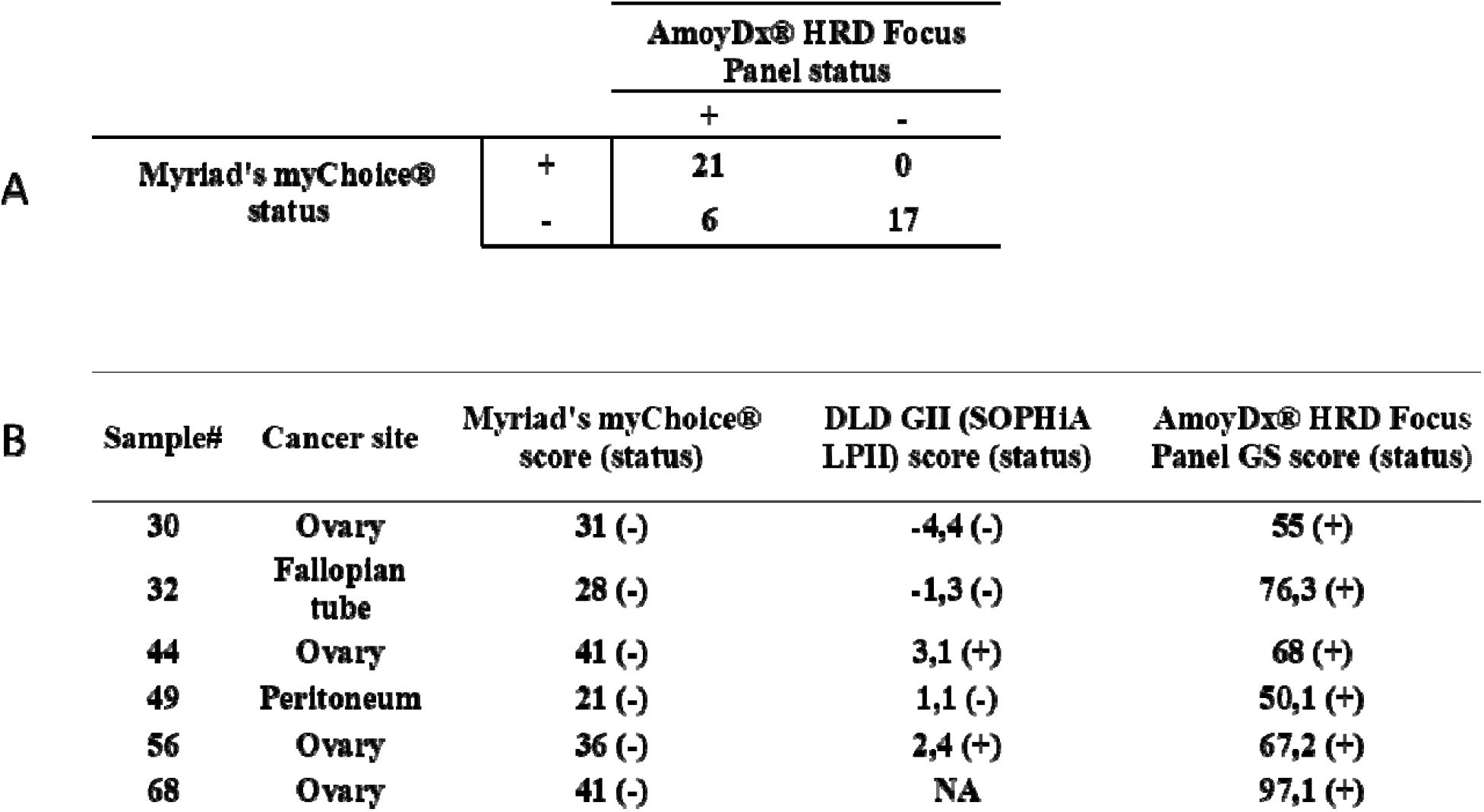
A) Contingency table of positive/negative samples showing concordance between Myriad’s myChoice and AmoyDx® solution. Note excess of false positives with complete sensitivity. B)Detailed information on the 6 samples with categorical divergencies between Myriad’s myChoice and AmoyDx® solution (with additional information of SOPHiA LPII GII, when available).

Four ovarian tumor samples, one fallopian tube and one peritoneum showed categorical discordances between Myriad’s myChoice® test and AmoyDx® HRD Focus Panel (Figure 6B). All samples with negative status based on Myriad’s myChoice® test, turned out to be the opposite status by AmoyDx® HRD Focus Panel, only one of them with borderline score. Interestingly AmoyDx® solution does not show the same linearity near the threshold of 42 (from Myriad’s myChoice®)as SOPHiA LPII solution does, which makes counseling near threshold more of a challenge. Moreover 2 out of the 3 discordant samples between SOPHiA LPII (positive) and Myriad’s myChoice® (negative) were actually concordant positive between SOPHiA LPII and AmoyDx®, serving again as caution when counselling samples with borderline results (Figure 6B).

## DISCUSSION

The results presented here show the power and robustness of HRD-One considering its ability to correctly discriminate positive and negative samples with regard to their HRD status. Using a validation set of 59 ovarian samples, an overall categorical concordance between Myriad’s myChoice® and HRD-One of 94.74% was observed. The analytical sensitivity of the test was estimated at 95.7%, specificity of 94.1%, PPV of 91.7% and NPV of 97%. If taken into consideration, both the uncertainty margin from the DLD GII score (0.2-3.2) and the average replica difference margin (1.62 ±0.88) it is clear that scores between 0 and +3.5 should be interpreted with due caution. Novel studies have suggested that even a lower threshold of 33 (equivalent to -1.5) showed improved survival after platinum monotherapy in high-grade serous ovarian cancer^4^. The threshold of 42 (+2.0) was established in a single study and further studies, that correlate not only *BRCA1/2* and genomic scars but also more data on treatment outcomes with PARPi, are needed to better understand for which patients and tumors PARPi is beneficial. With the current data we conclude and advise that for high-grade serous epithelial ovarian cancers an HRD-score of +2.0 (equivalent to 42 in myChoice®) should be used, with the cautionary recommendation that scores from +0.5 to +1.5 might be considered positive in tests performed at different dates or laboratories. In summary we have presented that our HRD-One score had more than 94% concordance with the current available commercial test and that if the 1.62-point variance margin would be taken into consideration the three samples that were discordant would fall into the uncertainty margin and all 59 samples could be classified as concordant. Therefore, we consider HRD-One a reliable alternative for HRD testing on ovarian samples where HRD status is expected to assist on the clinical management of patients. Moreover, even though AmoyDx® HRD Focus Panel has a high sensitivity to detect HRD status, in our validation it presented an excessive number of samples that failed quality control (7/44), excessive ‘false positive’ results (when compared to the other two solutions – 5/44) and lower linear or polynomial correlation with the other scores, which makes it, at the tested version, less optimal than the other solutions.

## Data Availability

Public data is depicted on the manuscript. Raw and identifiable data are held privately and under security by the testing laboratory according to local law.

## Ethics committee approval

**CAAE -55810222.0.0000.5455**

